# Incremental Clinical Value of Single-Molecule Nanopore Sequencing in Thalassemia Testing: A Prospective, Double-blind, Multicenter Study

**DOI:** 10.64898/2026.06.09.26354559

**Authors:** Yuancun Zhao, Haiyan Xu, Yan Chen, Haoqing Zhang, Jun He, Lihong Pang, Wenlan Liu, Jufang Tan, Zhenhai Fan, Jing Liu, Ling Shi, Lei Zhang, Xiangzhong Sun, Yamin Kong, Ruizhi Xiang, Ying Chen, Aiqi Cai, Jiale Xiang, Baosheng Zhu

## Abstract

**Background:** Thalassemia is one of the most common monogenic disorders worldwide. Current screening strategies combining hematological testing with molecular assays still carry a risk of missed diagnoses and suboptimal efficiency, particularly for complex structural variants and rare mutations.

**Methods:** In this prospective, double-blind, multicenter cohort study of 3,842 participants (3,362 pregnant women and 480 male partners), we conducted a head-to-head comparison to systematically evaluate the incremental clinical value and detection performance of single-molecule nanopore sequencing in thalassemia (SMITH) against conventional hematological testing and next-generation sequencing (NGS).

**Findings:** The overall concordance rate between NGS and SMITH was 98.6% (3789/3842). The discrepant cases (n=53) were directly attributed to the superior detection capabilities of SMITH, which successfully identified complex structural rearrangements—including 45 α-globin gene triplications and four HKαα alleles—that were missed by NGS. Furthermore, SMITH accurately detected four rare variants (αα^c.134_135insT^/αα, αα^c.-22(C>T)^/αα, β ^N^/β^c.316-290delinsAGGGCAATAATTT^, and β^3.5 kb deletion^/β^N^) and resolved ten *trans* and three *cis* configurations within the globin gene alleles. Clinically, these technical advantages translated to a 9.3% (5/54) increase in the detection rate of high-risk prenatal couples, effectively preventing one birth affected by moderate-to-severe thalassemia. Additionally, SMITH corrected a diagnostic discrepancy in one case (HKαα vs. -α^3.7^), sparing the couple from an unnecessary invasive procedure.

**Interpretation:** Our findings demonstrate that SMITH provides a powerful platform for resolving globin gene rearrangements, detecting rare variants, and enabling direct haplotype phasing. By effectively eliminating diagnostic blind spots, SMITH is expected to become an optimal method for thalassemia prevention programs.

**Funding:** This study was supported by the Chinese National Natural Science Foundation Projects 81760037 and 82271894.

**Research in context:** *Evidence before this study:* Next-generation sequencing (NGS) has substantially advanced carrier screening for thalassemia. However, its analytical blind spots—particularly in highly homologous regions, complex structural variants (SVs), and haplotype phasing—directly compromise the accuracy of genetic counseling and prenatal diagnosis. Long-read sequencing holds promise to address these gaps by generating reads that span entire gene clusters. Prior to this study, research on long-read sequencing for thalassemia screening had been extensive. To systematically evaluate its clinical value, we searched the literature published over the past decade. However, most previous studies compared long-read sequencing with conventional PCR-based methods, which do not allow assessment of its incremental value relative to current NGS-based screening pathways. To date, no large-scale, prospective, head-to-head study has compared NGS with single-molecule nanopore sequencing in a real-world prenatal screening setting. Therefore, the incremental clinical value of long-read sequencing in this context remains to be systematically quantified.

*Added value of this study:* In this prospective, double-blind, multicenter cohort study of 3,842 participants, we performed a head-to-head comparison between standard NGS and our novel Single-Molecule nanopore sequencing In THalassemia (SMITH) in a real-world prenatal screening setting. To our knowledge, this is the first large-scale, prospective study to directly compare long-read sequencing with standard NGS and to link technical performance with clinical outcomes. SMITH increased the detection rate of high-risk couples by 9.3%, prevented one moderate-to-severe thalassemia birth, and corrected a diagnostic misclassification that spared a couple from unnecessary invasive amniocentesis. These findings provide detailed, clinically actionable insights not previously reported.

*Implications of all the available evidence:* These findings establish SMITH as a major advance in thalassemia prevention. By eliminating key diagnostic blind spots—complex structural variants, rare mutations, and unresolved allelic phases—SMITH improves the accuracy of genetic counseling and prenatal diagnosis. The reduction in missed high-risk couples and preventable severe thalassemia births has direct implications for public health policy, particularly in high-prevalence regions. Future work should prioritise cost-effectiveness analyses and large-scale implementation studies to guide the integration of SMITH into routine population-based screening programmes.

## Introduction

Thalassemia is an autosomal recessive hemoglobinopathy caused by mutations in globin genes leading to an imbalance in globin chain synthesis.^1-3^ Globally, an estimated 56,000 individuals are born annually with clinically significant thalassemia, of whom approximately 30,000 require lifelong blood transfusions.^3^ In China, it is particularly prevalent in southern provinces, and carrier rates can reach up to 20%, posing a substantial burden on the healthcare system.^4^ Carrier screening and prenatal diagnosis have been successfully implemented to reduce the birth prevalence of severe thalassemia. Consequently, this effective approach has been established as a critical public health strategy.^5^

The current standard of care typically involves a two-step approach: initial hematological screening (complete blood count and hemoglobin electrophoresis) followed by genetic testing for positive individuals.^6,7^ At the genetic testing level, traditional techniques (gel electrophoresis, Gap-PCR, and RDB) demonstrate restricted detection spectra, being principally capable of identifying common pathogenic variants,^7^ and frequently fail to detect de novo mutations. Although next-generation sequencing (NGS) has markedly improved the detection of single-nucleotide variants and common copy-number variants compared with conventional PCR-based methods,^5,6,8,9^ NGS struggles with technical blind spots, including the detection of highly homologous *HBA* gene cluster and resolving complex structural variants (e.g., *HBA* triplications), and the phasing of variants without parental inheritance data. For α-thalassemia, the sensitivity of standard NGS can be as low as 55.8%.^10^ These technical constraints may lead to false-negative results and unnecessary invasive prenatal diagnostic procedures prompted by ambiguous genotype information. Consequently, there is an urgent clinical need for a comprehensive screening method to improve the accuracy of detection.

Long-read sequencing addresses residual clinical risks through its capacity to generate long reads spanning entire gene clusters.^11^ This technology has demonstrated superior performance in detecting thalassemia variants. This capability enables direct detection of complex structural rearrangements and determination of allelic phase in a single assay, eliminating the requirement for pedigree analysis.^1,12–14^ Long-read sequencing has proven clinically valid, increasing thalassemia variant detection by 19%.^15^ However, prior comparisons have focused on PCR-based panels,^1,12–14^ which do not allow for a comprehensive assessment of current NGS routine screening pathways. The only direct NGS versus long-read sequencing study had a small sample size (n=52) and lacked clinical follow-up.^13^ Thus, the true clinical superiority of long-read sequencing in real-world, particularly its efficacy as a component of thalassemia screening and its direct influence on pregnancy outcomes, remains inadequately investigated.

In this prospective, blinded, multicenter study, we enrolled 3,362 pregnant women and their male partners to perform a head-to-head comparison of Single-Molecule nanopore sequencing In THalassemia (SMITH) and NGS in a prenatal screening workflow. We focused on its ability to correct screening results and prevent the birth of moderate-severe thalassemia traits. By demonstrating the superior accuracy of SMITH in identifying carriers and preventing unnecessary invasive diagnosis, this study provides evidence to enhance the precision of genetic counseling and prenatal diagnosis, while also offering a reference for real-world large-scale population prenatal thalassemia screening.

## Materials and Methods

### Study Participants and Ethical Approval

Blood samples were prospectively collected from 3,842 individuals (3,362 pregnant women and 480 male partners) presenting for prenatal thalassemia screening. All pregnant women concurrently underwent both NGS and SMITH. Male partners of women confirmed as carriers by genotyping subsequently underwent the identical testing protocol (Fig. 1). Participants were included in the study based on the following criteria: (1) pregnant women had undergone both standard hematological screening and hemoglobin electrophoresis; (2) gestational age was ≤22 weeks; and (3) written informed consent was obtained from all subjects. Identified high-risk couples were followed prospectively to collect prenatal diagnostic outcomes. The First People’s Hospital of Yunnan Province served as the principal coordinating center. The study protocol was approved by the Institutional Review Board of the First People’s Hospital of Yunnan Province (Approval No. KHLL2024-KY259) and the Office of China Human Genetic Resources Administration (Approval No. [2025] CJ0040). The five other participating centers obtained corresponding ethical approvals from their institutional review boards.

**Figure 1.**
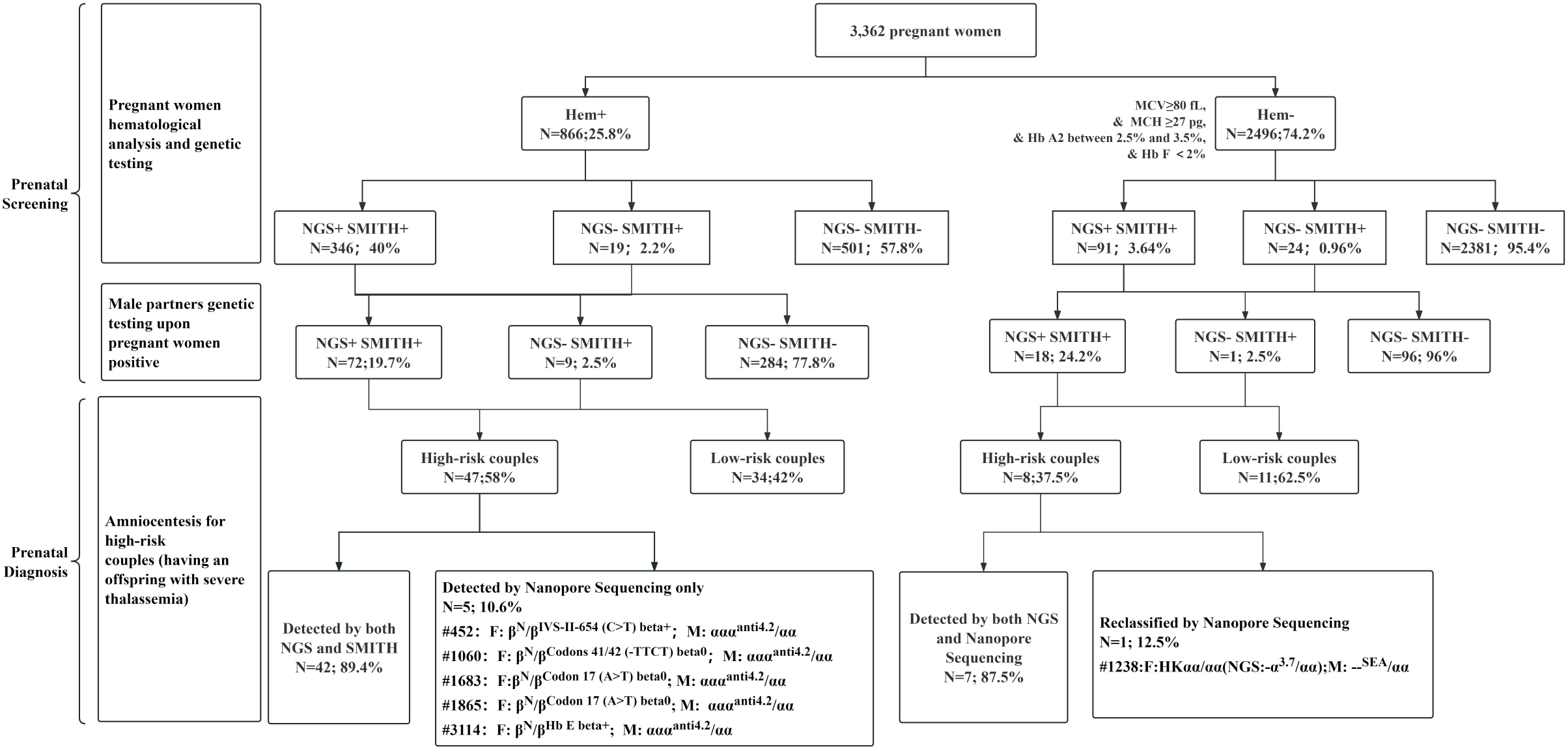
Thalassemia Prenatal Carrier Screening Study Flowchart. The diagram outlines the sequential screening strategy applied to our cohort. All pregnant women first underwent hematological analysis and then proceeded to NGS and SMITH. When pregnant women tested positive, their spouses were subsequently tested. Through this strategy, single-molecule nanopore sequencing identified five extra high-risk couples and corrected the screening result for one couple, avoiding unnecessary prenatal diagnosis. Abbreviations: MCV, mean corpuscular volume; MCH, mean corpuscular hemoglobin; Hb A2, hemoglobin A2; Hb F, hemoglobin F; NGS, next-generation sequencing; SMITH, single-molecule nanopore sequencing in thalassemia; Hem+, hematologically positive; Hem-, hematologically negative; N+, NGS positive; N-, NGS negative; SMITH+, SMITH positive; SMITH-, SMITH negative; F, Female; M, Male.

### Hematological Analysis

Hematological parameters were determined using an automated hematology analyzer (Sysmex, Japan). Hemoglobin analysis was performed utilizing either high-performance liquid chromatography (Bio-Rad, USA) or capillary electrophoresis (Sebia, France; Helena, USA). The reference intervals indicating a normal hematological profile were defined as follows: mean corpuscular volume (MCV) ≥80 fL, mean corpuscular hemoglobin (MCH) ≥27 pg, hemoglobin A2 (Hb A2) levels between 2.5% and 3.5%, and hemoglobin F (Hb F) ≤2%.

### Single-Molecule Nanopore Sequencing in Thalassemia

Samples were processed at the Beijing Genomics Institute, where SMITH was performed using the CycloneSEQ platform and the associated Cyclone library preparation kit (CycloneSEQ, Hangzhou, China). Briefly, genomic DNA was subjected to multiplex long-range PCR utilizing optimized primers to generate specific amplicons encompassing known structural variant (SV) regions, single-nucleotide variants (SNVs), and insertions/deletions (indels) within the *HBA1, HBA2*, and *HBB* genes. Following purification and end-repair, motor protein adapters were ligated using the Cyclone ligation module. In accordance with the manufacturer’s protocol, the prepared libraries were loaded onto a Cyclone sequencing chip using the provided anchoring and sequencing reagents, and sequencing was executed on a CycloneSEQ G100-ER sequencer.^14,16^

Raw FASTQ data were filtered, demultiplexed, and aligned to the human reference genome (hg38) using Minimap2 (version 2.26-r1175). SVs and SNVs were called using Sniffles (version 2.2), CuteSV (version 2.0.3), and FreeBayes (version 1.3.6). To resolve complex α-globin rearrangements, sequencing data were aligned against pre-designed multi-copy virtual genomes, enabling the accurate identification of complex SVs based on precise breakpoint positions. Subsequently, direct haplotype phasing was achieved utilizing LongPhase (v1.7) by integrating the combined SV and SNV data. All variants were annotated against the IthaNet and HbVar databases and visually confirmed using the Integrative Genomics Viewer (IGV). The detailed bioinformatics pipeline followed previously described protocols.^17^

### Thalassemia Carrier Screening by NGS

Parallel NGS evaluation was conducted at the Beijing Genomics Institute utilizing the MGISEQ-2000 platform (MGI, Shenzhen, China). Target sequences of the HBA1, HBA2, and HBB genes were first amplified and enriched via multiplex PCR. Sequencing libraries were then constructed according to the standard MGISEQ-2000 protocol, and paired-end 100 bp sequencing was performed on an MGISEQ-2000 chip. Finally, SNVs, SVs, and indels were called and annotated referencing the HbVar and IthaNet databases. The detailed bioinformatics pipeline for identifying hemoglobin gene mutations followed previously described protocols.^5,6^

### Confirmation of Discordant Variants

Discordant results between SMITH and NGS were orthogonally validated. α-globin gene triplications identified exclusively by SMITH were confirmed using a specifically designed Gap-PCR assay amplifying the anti3.7 and anti4.2 segments, as previously reported.^18^ Discordant SNVs and indels between the two sequencing platforms were verified via standard Sanger sequencing.

## Results

### Evaluation of the Performance of SMITH in Prenatal Screening

From June to October 2025, we enrolled 3,362 pregnant women and their male partners from six medical centers across southern China to conduct a prospective, blinded, multi-center parallel comparison (Fig. 1). All 3,362 pregnant women initially underwent standard hematological testing, which classified 866 (25.8%) as hematologically positive (Hem+) and 2,496 (74.2%) as hematologically negative (Hem-)(Fig. 1). Concurrently, all pregnant women underwent genetic screening using both NGS and SMITH.

Within the Hem+ cohort, 346 individuals (40%) were identified as carriers by both NGS and SMITH (NGS+SMITH+), while 501 (57.8%) were negative by both methods (NGS-SMITH-). Importantly, 19 discrepant cases (2.2%) were negative by NGS but positive by SMITH (NGS-SMITH+). In these cases, SMITH successfully detected complex structural variants (e.g., ααα^anti4.2^/αα and ααα^anti3.7^/αα) that were entirely missed by NGS. Similarly, in the Hem-cohort, while the vast majority (2,381, 95.4%) were concordantly negative, genetic testing still identified 91 (3.64%) NGS+SMITH+ cases and 24 (0.96%) NGS-SMITH+ cases (Fig. 1).

Spousal genetic testing was subsequently performed for the partners of carrier women to determine the reproductive risk. In the Hem+ arm, spousal testing ultimately identified 47 high-risk couples recommending prenatal diagnosis, and 34 low-risk couples. In the Hem-arm, spousal testing identified an additional eight prenatal high-risk couples and 11 low-risk couples.

Ultimately, this sequential screening strategy highlighted the substantial incremental clinical value of SMITH. Notably, SMITH uniquely identified five high-risk couples requiring prenatal diagnosis that had been missed by conventional NGS workflows. Furthermore, SMITH corrected a diagnostic error in one couple initially misdiagnosed by NGS; by accurately identifying a -α3.7 deletion as an HKαα allele, SMITH reclassified the couple as low-risk, thereby avoiding an unnecessary invasive prenatal diagnostic procedure (Fig. 1).

### SMITH Addresses Residual Gaps in Traditional Thalassemia Prenatal Carrier Screening

In this prospective cohort study, the clinical efficacy of SMITH was systematically evaluated through a blind comparison of two screening strategies: conventional screening strategies (hematological analysis with NGS) versus hematological analysis with SMITH. According to confirmed results, 54% (54/100) of couples were at high risk of delivering offspring with intermediate or major thalassemia (Table 1). All prenatal diagnostic outcomes and corresponding pregnancy decisions were rigorously documented (Supplemental Table 2).

**Table 1.** Detection Performance of Different Screening Strategies for Identifying High-Risk Couples. Among 100 carrier couples, 54 were confirmed as high-risk by traditional PCR-based method. Detection rates and 95% confidence intervals (CI) were calculated for each strategy.

| Screening Strategy | High-Risk<br>Couples(n) | Low-Risk<br>Couples(n) | Misclassified<br>Couples(n) | Detection<br>Rate (%) | 95%<br>CI (%) |
| --- | --- | --- | --- | --- | --- |
| Hematological Analysis | 47 | 53 | 0 | 87.0 | 75.6 – 93.6 |
| Hematological Analysis+NGS | 49 | 50 | 1 | 90.7 | 80.1–96.0 |
| Hematological Analysis+SMITH | 54 | 46 | 0 | 100.0 | 93.4 – 100.0 |

**Table 2.**
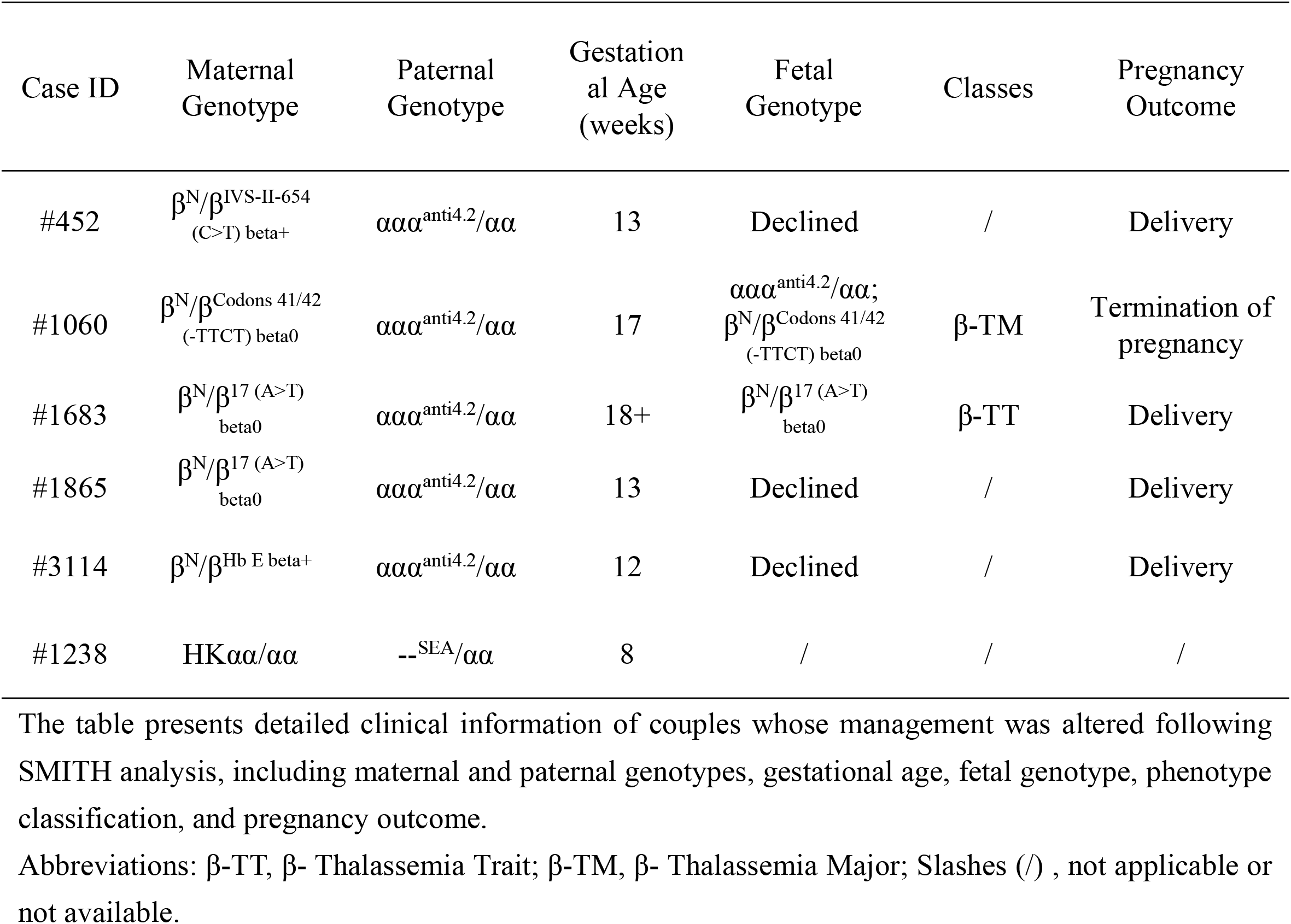
Summary of Prenatal Diagnoses and Outcomes with Altered Clinical Management Identified by SMITH. This table presents detailed clinical information of couples whose management was altered following SMITH analysis, including maternal and paternal genotypes, gestational age, fetal genotype, phenotype classification, and pregnancy outcome. Abbreviations: β-TT, β-Thalassemia Trait; β-TM, β-Thalassemia Major; Slashes (/), not applicable or not available.

Among 54 confirmed high-risk couples, hematological analysis detected only 47 (87.0%; 95% confidence interval [CI]: 75.6-93.6%) couples, leaving 13% undetected. Hematological analysis combined with NGS screening detected only 49 (90.7%; 95% CI: 80.1-96.0%) couples, leaving 9.3% undetected. These two strategies showed suboptimal sensitivity for thalassemia. In contrast, integration of SMITH with hematological analysis correctly identified all 54 high-risk couples, achieving 100% sensitivity (95% CI: 93.4–100.0%) (Table 1), effectively eliminating the residual gap in detection left by NGS-based screening. Further analysis revealed that all five cases (#452, #1060, #1683, #1865, and #3114) missed by NGS were involved with one αααanti4.2 carrier and one β-SNV carrier (β^N^/β^IVS-II-654 (C>T) beta+^, β^N^/β^17 (A>T) beta0^, β^N^/β^Hb E beta+^, β^N^/β^Codons 41/42 (-TTCT) beta0^) (Fig. 1 & Table 2). The dosage effect of the α-globin gene triplication disrupts hemoglobin synthesis balance, exacerbating the clinical severity of β-thalassemia.^19–21^ In these five couples, although three declined prenatal diagnosis and were lost to follow-up, the integration of SMITH into the screening pathway still enabled the successful prevention of one birth affected by moderate-to-severe thalassemia,^22^ clearly demonstrating its clinical utility in reducing the burden of hemoglobinopathies (Fig. 1 & Table 2).

### SMITH Reveals Hidden Risks in Conventional Screening Strategies

By virtue of its high-resolution, long-read capabilities, SMITH accurately resolved 34 cases of maternal α-globin triplications that had eluded detection by both NGS and routine hematological analysis (groups A–E). Among the male partners of these women, three tested positive. In two families (groups D and E), the partners also carried α-triplications, while in a third family (group C), the partner carried a structural hemoglobin variant (Hb Hamilton), a combination that confers no risk of severe thalassemia in the offspring (Supplemental Table 2). Although these specific diagnostic omissions did not result in adverse clinical consequences within our cohort, they highlight a critical systemic flaw: under the standard NGS workflow, a missed maternal triplication fails to trigger partner testing. If the untested partner happened to carry a β-thalassemia variant, this screening blind spot could leave the couple’s risk unrecognized, potentially leading to the preventable birth of a child with severe thalassemia.

Furthermore, four pregnant women in our cohort (group F) exhibited discordant genotyping results between the two platforms, with NGS misclassifying HKαα cases as -α^3.7^ deletions. Among their male partners, only one tested positive, carrying the --^SEA^ deletion (group G) (Fig. 1 & Table 2 & Supplemental Table 2). Based solely on the NGS findings, this specific parental genotype combination (-α^3.7^/--^SEA^) would have mandated a recommendation for prenatal diagnosis. However, SMITH analysis provided a timely and accurate genotypic revision to HKαα and --^SEA^, which is a low-risk combination. This correction carried significant clinical implications, successfully averting an unnecessary invasive diagnostic procedure and alleviating both the psychological distress and financial burden for the affected family.

### Comprehensive Detection of Complex Rearrangements, Rare Variants, and Allelic Phase by SMITH

Building on the above clinical findings, we further investigated the molecular basis underpinning the performance of SMITH. Within our cohort, the concordance rate between NGS and SMITH was 98.6% (3,789/3,842), with a discordance rate of 1.4% (53/3,842). Although this 1.4% reflects a numerically modest incremental diagnostic yield, SMITH achieved 100% detection sensitivity for complex structural rearrangements, successfully identifying 45 α-globin gene triplications (αααanti3.7 and αααanti4.2) and four HKαα alleles that were missed by standard NGS (Fig. 2A-C & Supplemental Table 3). Orthogonal validation using conventional Gap-PCR demonstrated complete concordance with the SMITH results for all 49 of these complex samples.

**Figure 2.**
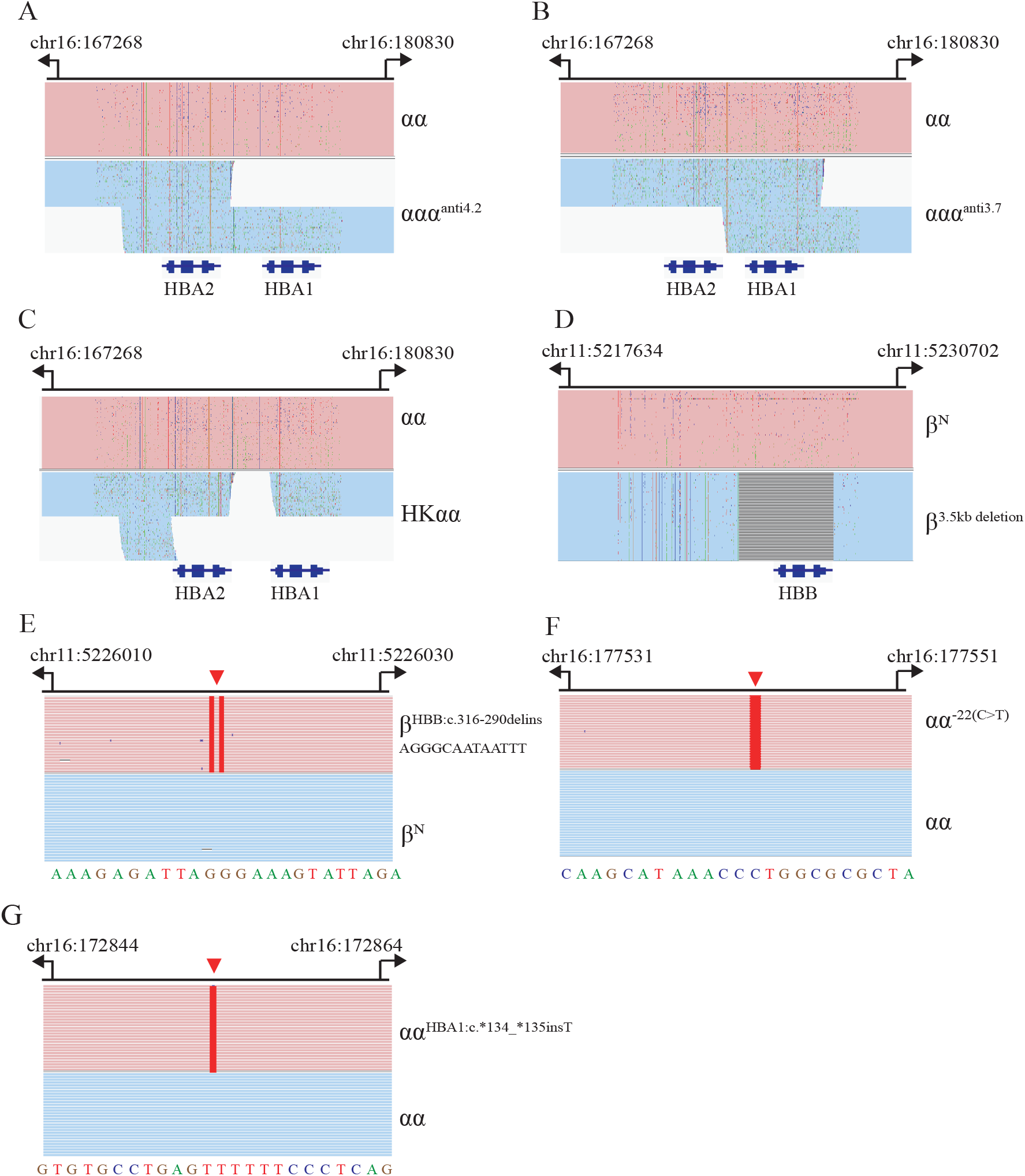
Integrative Genomics Viewer (IGV) Plots of Variants Identified Exclusively by SMITH. Both the normal and variant alleles are visible in each profile. (A-C) Complex structural rearrangements: (A) ααα^anti4.2^/αα; (B) ααα^anti3.7^/αα; (C) HKαα/αα; (D-G) Rare variants: (D) β^3.5 kb deletion^/β^N^; (E) β^N^/β^c.316-290delinsAGGGCAATAATTT^; (F) αα^-22 (C>T)^/αα; (G) αα^HBA1: c.*134_*135insT^/αα

Furthermore, SMITH successfully identified four rare variants within the α- and β-globin gene clusters that fell outside the detection scope of NGS. These included two variants in the *HBA* gene (αα^c.134_135insT^/αα and αα^-22 (C>T)^/αα), and two variants in the *HBB* gene (β^N^/β^c.316-290delinsAGGGCAATAATTT^ and β^3.5 kb deletion^/β^N^) (Fig. 2D-G & Supplemental Table 3). All four rare variants were subsequently and independently validated via Sanger sequencing.

Accurate reproductive risk assessment necessitates not only the detection of variants but also the precise determination of their allelic configuration (phasing). Within our cohort, SMITH successfully resolved ten *trans* (Supplemental Fig. 1A–H & Supplemental Table 4) and three *cis* (Supplemental Fig. 1I– J & Supplemental Table 4) configurations directly from maternal sequencing data. This capability provided critical phasing information for genetic counseling that conventional NGS alone could not deliver without additional pedigree analysis.

### Hematological Parameters of α-globin Triplication Combined with β-SNV

The gene dosage effect of multi-copy variants modulates hematological phenotypes and complicates prenatal diagnostic interpretation. Consistent with previous reports,^19,21,23^ isolated carriers of α-globin triplications (including αααanti3.7 and αααanti4.2) exhibited normal hematological parameters (Supplemental Fig. 2A). However, co-inheritance with β-thalassemia mutations had a marked clinical impact.^19–21^ In our cohort, two transfusion-independent cases were identified as compound heterozygotes with hematological screening results (Supplemental Table 3). While their hematological indices were abnormal compared to carriers of isolated α-globin triplications, they did not differ significantly from those of simple β-thalassemia heterozygotes (Supplemental Fig. 2B).

## Discussion

Prior assessments of long-read sequencing have primarily focused on technical performance against PCR-based panels,^1,12,13,15^ leaving its incremental value over current NGS pathways and its direct impact on pregnancy outcomes largely unexplored in the real world. To bridge this gap, we conducted the first prospective, multicenter, parallel-controlled study to perform a head-to-head comparison of SMITH and conventional NGS in a real-world prenatal carrier screening setting for thalassemia.

Our findings demonstrate that SMITH significantly enhanced the detection rate of high-risk couples by 9.3%, thereby directly preventing a birth affected by moderate-to-severe thalassemia.^22^ Furthermore, SMITH successfully rectified a misdiagnosed case, sparing the couple from an unnecessary invasive prenatal diagnosis. This incremental clinical value is directly attributable to the superior resolution of SMITH for complex structural rearrangements and highly homologous regions. In our cohort, SMITH achieved 100% sensitivity for detecting α-globin gene triplications, all of which were challenging to detect using conventional short-read sequencing methods. These findings demonstrate that SMITH provides a powerful approach for resolving highly homologous regions, precisely defining recombination breakpoints, and accurately characterizing complex structural rearrangements in the α-globin gene cluster,^24–26^ delivering significant clinical value for risk assessment and reproductive decision-making.

The ability of SMITH to accurately detect α-globin triplications addresses a critical need in regions with high β-thalassemia prevalence. In southern China, β-thalassemia carrier rates range from 2.34% to 7.05% in Guangxi province,^4,5^ with α-triplication frequencies of 0.92–2.25%.^5^ Co-inheritance of α-triplication exacerbates α/β-globin chain imbalance, increasing β-thalassemia severity.^19–21^ Given its clinical impact and population frequency, expert consensus now recommends that β-thalassemia genotyping be routinely performed for the partner of any individual identified as an α-triplication carrier.^27^ Compared to NGS, SMITH identified 34 α-globin gene triplications in pregnant women, with all male partners either testing negative or carrying an α-thalassemia genotype. Although these screening omissions in the NGS screening strategy were without clinical sequelae, this blind spot poses a hidden threat: when the untested paternal allele carries a β-SNV, the couple’s combined risk for having a child with severe thalassemia goes unrecognized, leading to preventable affected births. Consequently, the current workflow conditions paternal analysis on maternal test results and is further limited by incomplete variant detection. This approach fails to proactively identify high-risk couples, leaving a preventable gap in thalassemia prevention.

Notably, even with the precise identification of α-triplication carriers by SMITH, clinical management remains challenging. The phenotypic expression of α-triplication co-inherited with β-globin variants is notably heterogeneous.^23,28–30^ In our cohort, two affected individuals showed hematological parameters comparable to those with isolated β-globin variants and required no transfusion. This heterogeneity underscores the need for large-scale studies to clarify the clinical significance of α-triplication. Importantly, given this heterogeneity, clinicians should exercise caution in prenatal diagnostic decision-making, integrating both genotypic and phenotypic data to guide appropriate clinical management.

SMITH enables reliable linkage of distantly spaced variants to individual DNA molecules. This allows direct analysis of complete haplotypes containing multiple variants without the need for pedigree information, facilitating precise discrimination of *cis/trans* configurations. In our cohort, SMITH accurately resolved ten trans and three cis configurations, providing critical guidance for genetic counseling. This capability is particularly essential for accurate risk assessment in compound heterozygotes, while streamlining the clinical workflow by eliminating the need for time-intensive family segregation studies.

SMITH achieves comprehensive amplification of all targeted sequences within primer-defined regions in a single reaction, demonstrating expanded detection coverage and enhanced identification of variants typically missed by conventional PCR panel^25^ and NGS.^13^ Among 3,842 individuals, 53 (1.4%) showed discordant results between NGS and SMITH, with SMITH additionally detecting four rare variants. However, due to limitations in PCR enzyme processivity, SMITH currently cannot achieve full-length amplification of the HBA and HBB genes, consequently restricting detection of variants outside primer-covered regions. Technological advancements will unveil the molecular pathogenesis in cases with unexplained anemia. Furthermore, new technological evolution introduces more challenges in variant interpretation, particularly in accurately determining the clinical significance of rare variants, emphasizing the urgent need for standardized interpretation frameworks in thalassemia counseling.

With progressive cost reduction, long-read sequencing, characterized by extensive detection range and high accuracy, emerges as a promising new standard for population-scale thalassemia screening.^25^ Operationally, this technology supports single-sample independent testing, reducing average reporting time by approximately 50% compared to NGS, minimizing processing delays and resource inefficiencies inherent to multi-step testing paradigms.^31^ In contrast to the bulky and costly PacBio Sequel II system, the compact, benchtop design of the nanopore sequencing platform facilitates on-site thalassemia testing.^26^ By integrating the sequencer, reagents, and software systems within the laboratories of collaborating medical institutions, both sample transportation time and associated costs were substantially reduced, improving testing efficiency. This model is particularly well-suited for regional prevention and control programs for genetic disorders such as thalassemia, providing a practical pathway for technology deployment at the grassroots level and enabling large-scale precision screening.

In conclusion, this large-scale, real-world multicenter study establishes the superior performance of SMITH over short-read sequencing, particularly for resolving complex structural variants. Given that inherent limitations of traditional methods can result in misclassification of carrier status and the subsequent birth of children with severe thalassemia,^11^ the comprehensive genotyping capability of SMITH is essential for accurate genetic risk assessment and precision screening.^25^ The adoption of such high-resolution technology not only provides robust support for reproductive decision-making and prevents the birth of children affected by thalassemia, but also drives a fundamental paradigm shift from multi-technology screening to a streamlined, one-step precision diagnostic model.

## Data Availability

All data produced in the present study are available upon reasonable request to the authors
All data produced in the present work are contained in the manuscript

## Contributors

BS Zhu, JL Xiang, Y Chen, HQ Zhang, J He, WL Liu, LH Pang, YC Zhao, and HY Xu conceived and designed the study. YC Zhao, HY Xu, RZ Xiang, and Y Chen performed the experiments. HY Xu, YC Zhao, JF Tan, ZH Fan, J Liu, L Shi, and L Zhang collected the samples. HY Xu, JL Xiang, XZ Sun, RZ Xiang, and Y Chen wrote the manuscript. All authors accessed and verified the data. All authors reviewed and approved the final version.

